## Supplementary File 1. PRISMA 2009 Checklist and GATHER checklist for "Model-based spatial-temporal mapping of opisthorchiasis in endemic countries of Southeast Asia"

Supplementary File 1A. PRISMA 2009 Checklist

| Section/topic | # | Checklist item | Reported on page # |
|---|---|---|---|
| **TITLE** | | | |
| Title | 1 | Identify the report as a systematic review, meta-analysis, or both. | 1 |
| **ABSTRACT** | | | |
| Structured summary | 2 | Provide a structured summary including, as applicable: background; objectives; data sources; study eligibility criteria, participants, and interventions; study appraisal and synthesis methods; results; limitations; conclusions and implications of key findings; systematic review registration number. | 2 |
| **INTRODUCTION** | | | |
| Rationale | 3 | Describe the rationale for the review in the context of what is already known. | 2-5 |
| Objectives | 4 | Provide an explicit statement of questions being addressed with reference to participants, interventions, comparisons, outcomes, and study design (PICOS). | 2-5 |
| **METHODS** | | | |
| Protocol and registration | 5 | Indicate if a review protocol exists, if and where it can be accessed (e.g., Web address), and, if available, provide registration information including registration number. | 17; Figure 1-figure supplement 1 |
| Eligibility criteria | 6 | Specify study characteristics (e.g., PICOS, length of follow-up) and report characteristics (e.g., years considered, language, publication status) used as criteria for eligibility, giving rationale. | 17-18; Figure 1-figure supplement 1 |
| Information sources | 7 | Describe all information sources (e.g., databases with dates of coverage, contact with study authors to identify additional studies) in the search and date last searched. | 17; Figure 1-figure supplement 1 |
| Search | 8 | Present full electronic search strategy for at least one database, including any limits used, such that it could be repeated. | 17; Figure 1-figure supplement 1 |
| Study selection | 9 | State the process for selecting studies (i.e., screening, eligibility, included in systematic review, and, if applicable, included in the meta-analysis). | 17-18; Figure 1-figure supplement 1 |
| Data collection process | 10 | Describe method of data extraction from reports (e.g., piloted forms, independently, in duplicate) and any processes for obtaining and confirming data from investigators. | 18; Figure 1-figure supplement 1 |
| Data items | 11 | List and define all variables for which data were sought (e.g., PICOS, funding sources) and any assumptions and simplifications made. | 18; Figure 1-figure supplement 1 |
| Risk of bias in individual studies | 12 | Describe methods used for assessing risk of bias of individual studies (including specification of whether this was done at the study or outcome level), and how this information is to be used in any data synthesis. | Figure 2-figure supplement 1-source data 1 |
| Summary measures | 13 | State the principal summary measures (e.g., risk ratio, difference in means). | 20-23 |
| Synthesis of results | 14 | Describe the methods of handling data and combining results of studies, if done, including measures of consistency (e.g., $I^2$) for each meta-analysis. | 20-23 |
| Risk of bias across studies | 15 | Specify any assessment of risk of bias that may affect the cumulative evidence (e.g., publication bias, selective reporting within studies). | Figure 2-figure supplement 1-source data 1 |
| Additional analyses | 16 | Describe methods of additional analyses (e.g., sensitivity or subgroup analyses, meta-regression), if done, indicating which were pre-specified. | 23-26 |

| Section/topic | # | Checklist item | Reported on page # |
|---|---|---|---|
| **RESULTS** | | | |
| Study selection | 17 | Give numbers of studies screened, assessed for eligibility, and included in the review, with reasons for exclusions at each stage, ideally with a flow diagram. | Figure 1 |
| Study characteristics | 18 | For each study, present characteristics for which data were extracted (e.g., study size, PICOS, follow-up period) and provide the citations. | Table1; Figure 1-2; Figure 2-source data 1; Figure 2-figure supplement 1-source data 1 |
| Risk of bias within studies | 19 | Present data on risk of bias of each study and, if available, any outcome level assessment (see item 12). | Figure 2-figure supplement 1-source data 1 |
| Results of individual studies | 20 | For all outcomes considered (benefits or harms), present, for each study: (a) simple summary data for each intervention group (b) effect estimates and confidence intervals, ideally with a forest plot. | N/A |
| Synthesis of results | 21 | Present results of each meta-analysis done, including confidence intervals and measures of consistency. | Table 2-3; Figure 3-6; Figure 6-figure supplement 1-9 |
| Risk of bias across studies | 22 | Present results of any assessment of risk of bias across studies (see Item 15). | Figure 2-figure supplement 1-source data 1 |
| Additional analysis | 23 | Give results of additional analyses, if done (e.g., sensitivity or subgroup analyses, meta-regression [see Item 16]). | 7; Figure 3-figure supplement 1; Figure 3-figure supplement 1-source data 1; Figure 2-source data 2 |
| **DISCUSSION** | | | |
| Summary of evidence | 24 | Summarize the main findings including the strength of evidence for each main outcome; consider their relevance to key groups (e.g., healthcare providers, users, and policy makers). | 5-9 |
| Limitations | 25 | Discuss limitations at study and outcome level (e.g., risk of bias), and at review-level (e.g., incomplete retrieval of identified research, reporting bias). | 13-16 |
| Conclusions | 26 | Provide a general interpretation of the results in the context of other evidence, and implications for future research. | 9-13 |
| **FUNDING** | | | |
| Funding | 27 | Describe sources of funding for the systematic review and other support (e.g., supply of data); role of funders for the systematic review. | 27 |

*From:* Moher D, Liberati A, Tetzlaff J, Altman DG, The PRISMA Group (2009). Preferred Reporting Items for Systematic Reviews and Meta-Analyses: The PRISMA Statement. PLoS Med 6(7): e1000097. doi:10.1371/journal.pmed1000097

For more information, visit: **www.prisma-statement.org**.

Supplementary File 1B. GATHER checklist of information that should be included in reports of global health estimates

| # | Checklist item | Reported on page # |
|---|---|---|
| **Objectives and funding** | | |
| 1 | Define the indicator(s), populations (including age, sex, and geographic entities), and time period(s) for which estimates were made. | 20-23 |
| 2 | List the funding sources for the work. | 27 |
| **Data inputs** | | |
| For all data inputs from multiple sources that are synthesized as part of the study: | | |
| 3 | Describe how the data were identified and how the data were accessed. | 17 |
| 4 | Specify the inclusion and exclusion criteria. Identify all ad-hoc exclusions. | 17-18; Figure 1-figure supplement 1 |
| 5 | Provide information about all included data sources and their main characteristics. For each data source used, report reference information or contact name/institution, population represented, data collection method, year(s) of data collection, sex and age range, diagnostic criteria or measurement method, and sample size, as relevant. | Table1; Figure 1-2; Figure 2-source data 1; Figure 2-figure supplement 1-source data 1 |
| 6 | Identify and describe any categories of input data that have potentially important biases (e.g., based on characteristics listed in item 5). | Table1; Figure 1; Figure 2-figure supplement 1-source data 1 |
| For data inputs that contribute to the analysis but were not synthesized as part of the study: | | |
| 7 | Describe and give sources for any other data inputs. | Figure 2-source data 1; Figure 2-figure supplement 1-source data 1 |
| For all data inputs: | | |
| 8 | Provide all data inputs in a file format from which data can be efficiently extracted (e.g., a spreadsheet rather than a PDF), including all relevant meta-data listed in item 5. For any data inputs that cannot be shared because of ethical or legal reasons, such as third-party ownership, provide a contact name or the name of the institution that retains the right to the data. | Figure 2-source data 1 |
| **Data analysis** | | |
| 9 | Provide a conceptual overview of the data analysis method. A diagram may be helpful. | 20-23 |
| 10 | Provide a detailed description of all steps of the analysis, including mathematical formulae. This description should cover, as relevant, data cleaning, data pre-processing, data adjustments and weighting of data sources, and mathematical or statistical model(s). | 19-23 |

| # | Checklist item | Reported on page # |
|---|---|---|
| 11 | Describe how candidate models were evaluated and how the final model(s) were selected. | 22 |
| 12 | Provide the results of an evaluation of model performance, if done, as well as the results of any relevant sensitivity analysis. | 23-24 |
| 13 | Describe methods of calculating uncertainty of the estimates. State which sources of uncertainty were, and were not, accounted for in the uncertainty analysis. | 23-26 |
| 14 | State how analytic or statistical source code used to generate estimates can be accessed. | 23 |
| **Results and discussion** | | |
| 15 | Provide published estimates in a file format from which data can be efficiently extracted. | Table 2-3; Figure 3-6; Figure 6-figure supplement 1-9; Source Data |
| 16 | Report a quantitative measure of the uncertainty of the estimates (e.g., uncertainty intervals). | Figure 4 |
| 17 | Interpret results in light of existing evidence. If updating a previous set of estimates, describe the reasons for changes in estimates. | 9-13 |
| 18 | Discuss limitations of the estimates. Include a discussion of any modelling assumptions or data limitations that affect interpretation of the estimates. | 13-16 |

*From:* Stevens GA, Alkema L, Black RE, Boerma JT, Collins GS, Ezzati M, Grove JT, Hogan DR, Hogan MC, Horton R, Lawn JE, Marušić A, Mathers CD, Murray CJL, Rudan I, Salomon JA, Simpson PJ, Vos T, Welch V. 2016. Guidelines for Accurate and Transparent Health Estimates Reporting: the GATHER statement. *PLOS Medicine* **13**:e1002116. doi:10.1371/journal.pmed.1002056
